# Evolution of Myocarditis Incidence at a Large Healthcare System Before and During COVID-19 Pandemic

**DOI:** 10.1101/2023.08.11.23294002

**Authors:** Brian C. Case, Ori Waksman, Hank Rappaport, Cheng Zhang, Ron Waksman

## Abstract

**Background:** Myocarditis is a recognized complication of severe acute respiratory syndrome coronavirus 2 (SARS-CoV-2) infection. Emerging studies further suggest an associated risk of myocarditis following administration of messenger RNA (mRNA) COVID-19 vaccinations. coronavirus disease 2019 (COVID-19) vaccinations. We investigated the incidence of myocarditis throughout the COVID-19 pandemic across a large healthcare system in the Washington, DC, metropolitan area.

**Methods:** This retrospective analysis of patients admitted from 2017-2022. Myocarditis cases were temporally divided into two cohorts based on year of admission (pre-pandemic 2017-2019; pandemic 2020-2022), which were compared for overall myocarditis incidence. The primary endpoint was in-hospital mortality.

**Results:** The cohort included 573 myocarditis patients (pre-pandemic=208, pandemic=365). From 2017-2019, the total number and rate of myocarditis cases was consistent. Overall cases of myocarditis increased during the pandemic (97, 126, 142 patients in 2020, 2021, 2022, respectively). Interestingly, the rate of myocarditis cases not related to COVID-19 or the vaccines stayed consistent (0.0674%, 0.0676%, 0.0807%), but the rate of myocarditis related to COVID-19 or the vaccines myocarditis increased each year (0.0210%, 0.0416%, 0.0480%). In-hospital mortality was similar between the two pre-pandemic and pandemic cohorts (5.35% versus 7.7%, 0.276).

**Conclusion:** Among hospitalized patients, during the COVID-19 pandemic, the incidence of myocarditis increased as compared to the pre-pandemic era. It appears this increase is associated with either the SARS-CoV-2 infection or COVID-19 vaccination. In-hospital outcomes did not differ during the pandemic, but ongoing research is needed to evaluate the long-term impact of myocarditis during the pandemic.

## Introduction

Evidence of myocardial injury as detected by elevated biomarkers or cardiac imaging is commonly observed among patients hospitalized with coronavirus 2019 disease (COVID-19). While several mechanisms have been identified, including stress cardiomyopathy, heart failure, and plaque destabilization, increased attention has been raised with regard to myocarditis, which has been reported even in asymptomatic severe acute respiratory syndrome coronavirus 2 (SARS-CoV-2) infection and the absence of previous cardiovascular disease^1-4^.

In addition to viral myocarditis, large retrospective studies and reports from the Vaccine Adverse Event Reporting System (VAERS) suggest significant association between myocarditis and administration of bivalent messenger RNA (mRNA) COVID-19 vaccinations^4, 5^. While cases of severe myocarditis with both viral infection and vaccination remain rare, the paucity of data regarding long-term cardiovascular outcomes has collectively challenged understanding of viral and immune-mediated myocarditis and has complicated vaccination efforts^3-6^.

Overall, there are scant data regarding the prevalence of myocarditis during the COVID-19 pandemic and the temporal effects of mRNA vaccinations on the community prevalence of myocarditis throughout the pandemic. In this analysis, we sought to investigate temporal trends between the incidence of myocarditis and different stages of the COVID-19 pandemic across a large healthcare system in the Washington, DC, metropolitan area.

## Methods

We analyzed the incidence of patients admitted into the hospital for myocarditis (based on International Classification of Diseases, Tenth Revision [ICD-10] code) in the MedStar Health system (11 hospitals in Washington, DC, and Maryland) from January 1, 2017, to December 31, 2022. Myocarditis was clinically diagnosed by the treating provider based on the elevation of troponin and/or imaging such as cardiac magnetic resonance imaging. The final diagnosis of myocarditis was made clinically by the treating provider according to their discretion. Patients with known infiltrative cardiomyopathy, i.e., sarcoidosis, were excluded from the analysis. The study was approved by the MedStar Health Institutional Review Board.

To evaluate the overall trend of myocarditis patients before and throughout the pandemic, overall cases were reported chronically over each year, and then compared from 2017 to 2019 (pre-COVID-19 pandemic) and 2020 to 2022 (during COVID-19 pandemic). The rate of myocarditis of a time period was estimated using the ratio of total myocarditis cases over the total number of admissions during that time period. The rate of myocarditis was evaluated based on an association with COVID-19 or the COVID-19 vaccine.

In terms of comparing the two cohorts, baseline patient characteristics were collected. In this analysis, the co-morbidities were identified using ICD-10 codes. Laboratory data, including inflammatory markers, were compared between the two groups. The primary clinical endpoint was in-hospital mortality, with a secondary endpoint of length of stay.

Categorical variables were expressed in counts and percentages. Continuous variables were expressed as mean ± SD. Categorical variables were compared using the χ^2^ test, and continuous variables were compared using Student’s t-test. A p-value below 0.05 was considered significant. All analysis were conducted using SAS 9.2 (SAS Institute, Cary, NC). The data that support the findings of this study are available from the corresponding author upon reasonable request.

## Results

The cohort included 573 myocarditis patients admitted between January 1, 2017, and December 31, 2022 (pre-pandemic: n=208, pandemic: n=365). There was a slightly higher proportion of males, with a mean age of 50.06 ± 20.13 years. There was evidence of racial disparity, with African Americans having a higher rate of myocarditis during the COVID-19 pandemic (59.7%) as compared to prior to the pandemic (47.2%). The reverse was seen in Caucasians (31.1% versus 45.7%). Rates of co-morbidities, including hypercholesterolemia, diabetes, and stroke, were higher in the COVID-19 pandemic cohort. A summary of baseline characteristics is shown in Table 1.

**Table 1:**
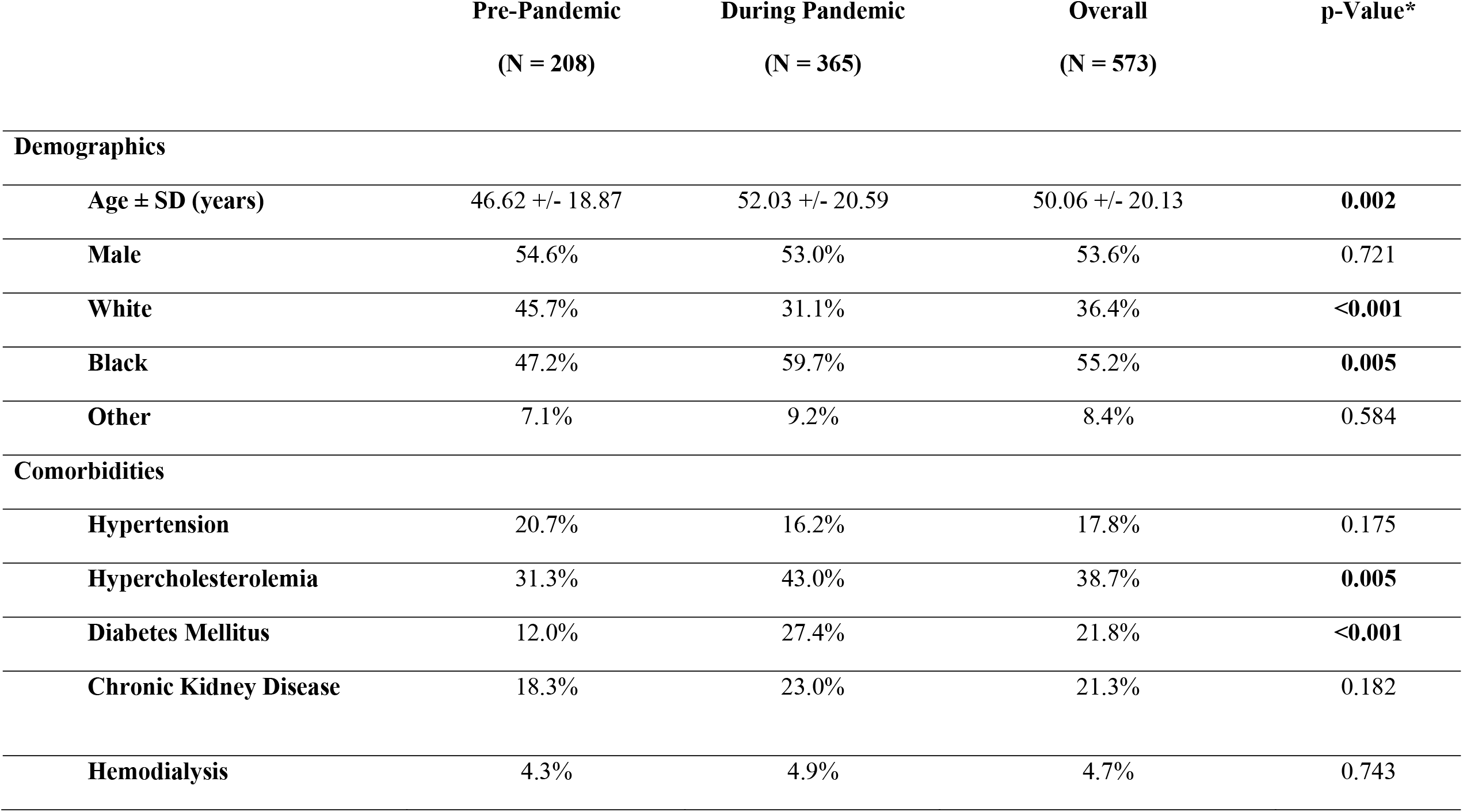
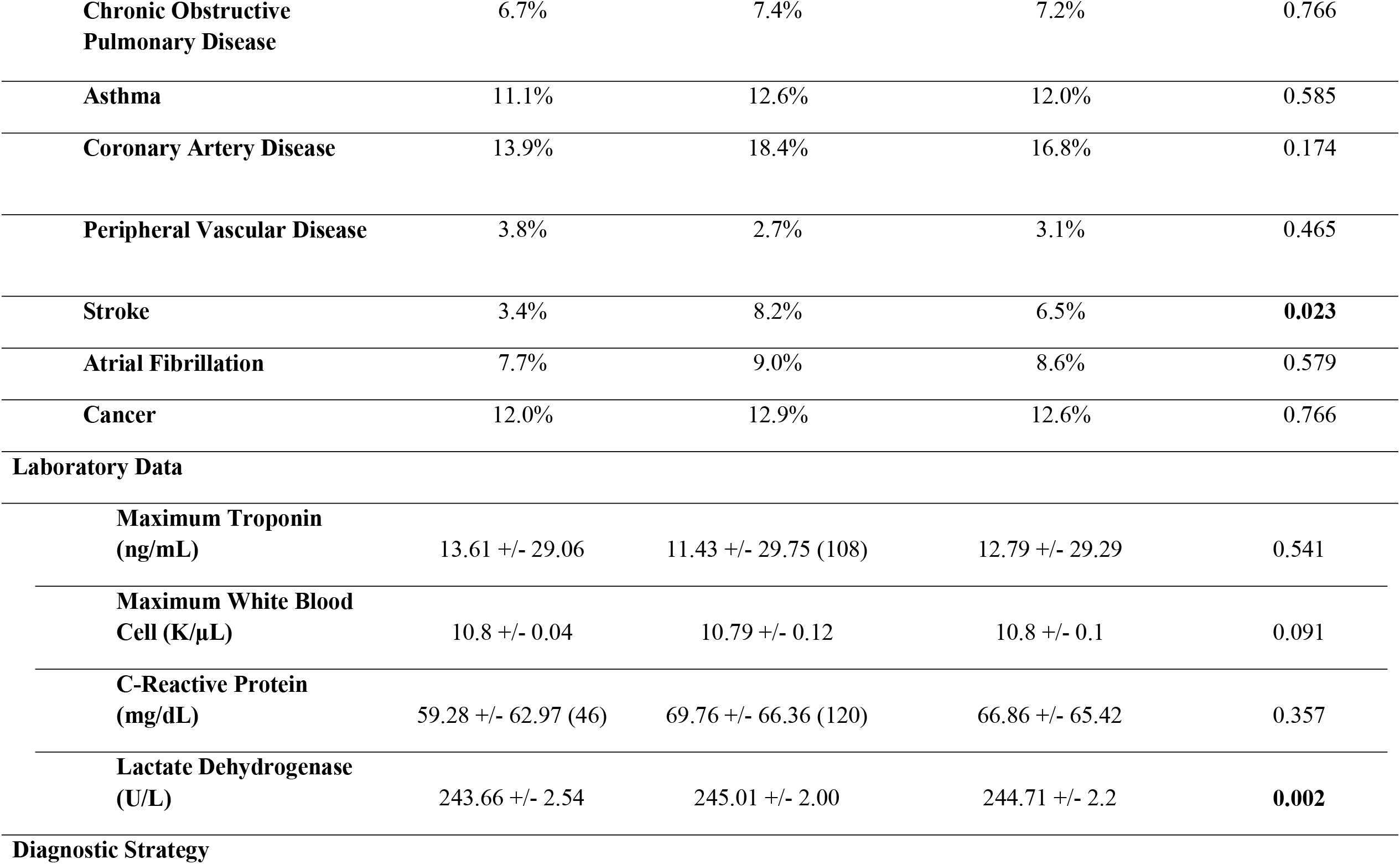
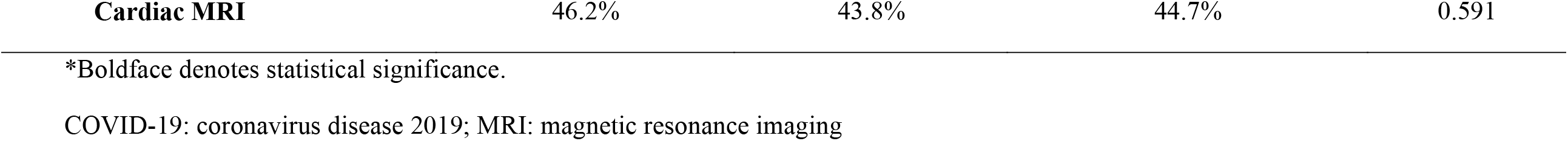
Baseline Characteristics, Laboratory Data, and Outcomes of Myocarditis Patients Before and During COVID-19 Pandemic.

In terms of diagnostic evaluation, white blood cell count, C-reactive protein, and maximum troponin were even between the two cohorts. Lactate dehydrogenase was slightly higher in the COVID-19 pandemic cohort. Finally, cardiac magnetic resonance imaging was conducted in 44.7% of our patient cohort overall. In terms of our clinical endpoints, in-hospital mortality was similar between the two arms (5.35% versus 7.7%, p=0.276) (**Table 2**). Further, length of stay was similar between the two cohorts.

**Table 2:** In-Hospital Outcomes of Myocarditis Patients Before and During COVID-19 Pandemic.

|  | **Pre-Pandemic**<br>**(N = 208)** | **During Pandemic**<br>**(N = 365)** | **Overall**<br>**(N = 573)** | **p-Value** |
| --- | --- | --- | --- | --- |
| **In-Hospital Mortality** | 5.3% | 7.7% | 6.8% | 0.276 |
| **Length of Stay** | 7.5 +/- 13.55 | 8.06 +/- 11.06 | 7.86 +/- 12.02 | 0.611 |

Figure 1. outlines the main finding of our analysis. From 2017 through 2019 (prepandemic) the total number, and rates (%), of myocarditis cases were fairly consistent. Starting in 2020 (start of pandemic), overall cases of myocarditis increased (97 patients) and continued to increase in 2021 (126 patients) and 2022 (142 patients). Interestingly, the rate of myocarditis cases not related to COVID-19 or the COVID-19 vaccine during the pandemic stayed consistent (0.0674%, 0.0676%, and 0.0807%). However, the rate of myocarditis cases related to previous exposure to COVID-19 or the COVID-19 vaccine increased each year (0.0210%, 0.0416%, and 0.0480%).

**Figure 1.**
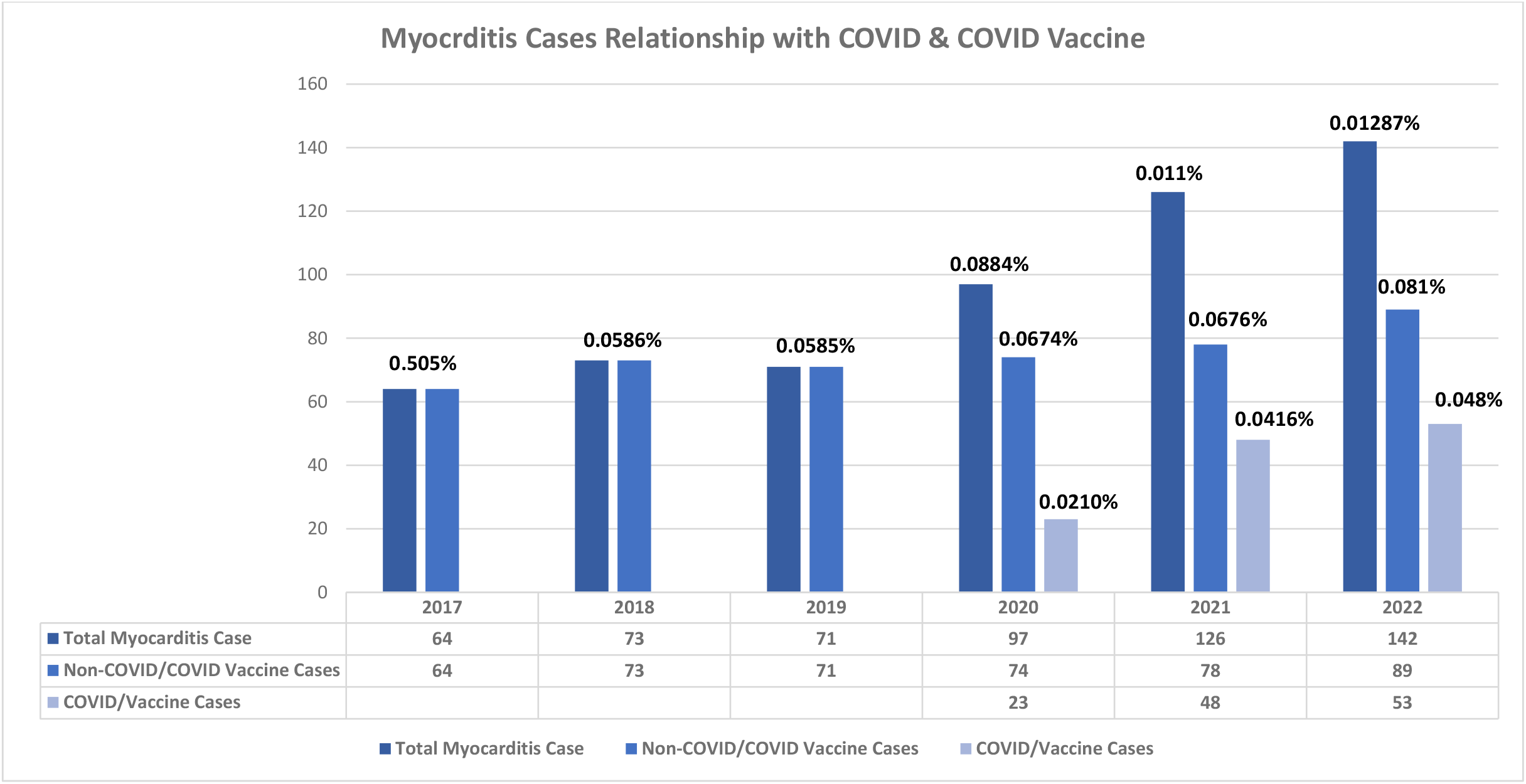
Overall Cases of Myocarditis Before and During COVID-19 Pandemic. % represents the rate of inpatient myocarditis cases of a year among the total inpatient admissions in that year.

## Discussion

In this multihospital retrospective cohort study of 573 patients hospitalized between 2017 and 2022 for myocarditis, we found that the community prevalence of myocarditis significantly increased between the pandemic years, 2020-2022, when compared to the pre-pandemic era, 2017-2019. Furthermore, between 2020 and 2022 the incidence of myocarditis admissions continued to increase each year, despite increased vaccination and the emergence of less-lethal SARS-CoV-2 variant strains. This increase in cases of myocarditis was chiefly seen in patients previously exposed to COVID-19 or the COVID-19 vaccine. Patients with myocarditis during the pandemic tended to be African American, with associated higher risk of diabetes, hypercholesterolemia, and history of stroke. Finally, in-hospital clinical outcomes did not differ pre-pandemic and during the pandemic.

Chronologically, the first COVID-19 case in Washington, DC, was reported on March 7, 2020, with the subsequent first wave, the alpha variant, peaking on April 28, 2020, with resolution by July 2020. The second wave, the delta variant, spread from December 2020 to February 2021, and the third wave, the omicron variant, spread from December 2021 to February 2022. The mortality rate decreased with each subsequent wave, with omicron being the most communicable but least lethal variant. In December 2020, the US Food and Drug Administration granted emergency use authorization to two formulations of (mRNA) COVID-19 vaccinations: two-shot-series Moderna mRNA-1273 and Pfizer-BioNTech BNT162b2^7^. The majority of Washington, DC, residents were administered the primary series by June 2021, with currently 99% of residents having received at least one dose and 81% receiving two doses of the primary series. (https://coronavirus.dc.gov/data/vaccination)

Our findings are surprising, as they suggest a sustained prevalence of myocarditis despite decreased community spread, decreased variant virulence, and increased vaccination trends. While significant national attention has been garnered by reports of vaccine-associated myocarditis, which suggest elevated myocarditis risk with sequential booster administration, the overall incidence of post vaccination myocarditis is estimated be to be 2.13 cases per 100,000 vaccinated persons and significantly lower than that of COVID-19 viral myocarditis^4, 5, 8^.

Furthermore, as it is mechanistically proposed that viral myocarditis is induced via a hyperinflammatory surge of cytokine and chemokine release in the setting of a maladaptive host immune response^9^, it is unclear why even with increased communicability, less virulent strains such as omicron would produce inflammation severe enough to induce myocardial injury. We hypothesize that sustained and rising prevalence of myocarditis is likely multifactorial and suggestive of sustained myocardial risk of less virulent variants, risk of sequential vaccinations, or potential sequalae of previous SARS-CoV-2 infection. Further investigation is warranted.

Limitations to our study include it being a retrospective analysis and diagnosis/treatment based on the treating physician. Further, association between myocarditis and SARS-CoV-2 infection or the COVID-19 vaccine is not fully determined. Using the ratio of total myocarditis cases over the total admissions to estimate the myocarditis rate counts the same patient twice or more and potentially underestimates the rate of myocarditis. Ideally, each patient is counted once after aggregating every patient’s chronic admission records. However, given that myocarditis is very rare, the absolute underestimation effect is almost negligible. Finally, prospective clinical outcomes were not measured.

In conclusion, during the COVID-19 pandemic, the rate of myocarditis inpatient admissions has increased, as compared to the pre-pandemic era, and remained sustained despite increased vaccination and less lethal strains. It appears that this increase is associated with either the SARS-CoV-2 infection or COVID-19 vaccination. In-hospital outcomes did not differ during the pandemic, but ongoing research is needed to evaluate the long-term impact of myocarditis during the pandemic.

## Data Availability

The data that support the findings of this study are available from the corresponding author upon reasonable request.

## ACKNOWLEDGEMENTS

Special acknowledgment to Jason Wermers, a paid medical writer employed by MedStar Health, for assistance in preparing this manuscript.

